# Limitations of Disease X vaccine efficacy and safety clinical trials

**DOI:** 10.1101/2025.10.11.25337791

**Authors:** Julien Flaig, Nicolas Houy

## Abstract

**Background:** In a vaccine clinical trial, the candidate is typically accepted or rejected based on predefined efficacy and safety thresholds. An efficacy threshold is the minimum measured treatment effect deemed statistically significant and clinically relevant. A safety threshold is the maximum number of adverse events deemed acceptable in the vaccine arm of the trial. However, the uncertainties and changing conditions met during the emergence of an unknown infectious disease (Disease X) may hinder such simple approaches.

**Methods:** We model the emergence of a Disease X with an SIR (susceptible-infectious-recovered) transmission model. A vaccine is available and the objective is to minimize the total cost over the course of the epidemic, which includes infection, vaccination, and adverse event costs. Uncertainties regarding transmission, vaccine, and cost parameters are represented by prior distributions. We simulate placebo-controlled efficacy and safety trials, and different vaccination policies. Under policies using trial results, the susceptible population is vaccinated if and only if the vaccine candidate passes both the efficacy and safety tests.

**Results:** In our baseline scenario and on average over uncertain parameters, using clinical trial outcomes to decide whether to vaccinate the population yields a lower expected total cost compared to indiscriminate emergency vaccination and to no intervention. However, testing both efficacy and safety yields a higher cost compared to testing safety alone. The difference counts in billions of dollars for a population of 10^8^ individuals. In our scenario, there is an optimal intermediate safety threshold value that best discriminates vaccine candidates. By contrast, there is no optimal intermediate efficacy threshold value. The incidence difference between the treated and control arms is even a misleading measure of vaccine performance.

**Conclusions:** We show with an example that simple threshold-based decision rules may not be appropriate to discriminate candidates in a Disease X vaccine clinical trial. Uncertainties and the disease dynamics need to be fully taken into account, which may require more advanced pattern recognition methods.

## 1 Introduction

Disease X is a hypothetical yet unknown disease with epidemic potential. It has been included in WHO’s list of priority diseases for research and development since 2018 [1, 2]. Responding in a timely manner to a Disease X outbreak is challenging due to the potentially fast dynamics of the epidemic and pervasive uncertainties – whether about the pathogen and its transmission, or about prospective interventions. This also applies to emerging and reemerging diseases. Vaccine development in such scenarios has received notable attention following the 2013-2016 Ebola crisis [3, 4], and since then with the development and approval of COVID-19 vaccines at an unprecedented speed [5] and a continued aim at further acceleration [6, 7].

Vaccine candidates are approved for use based on the assessment of their overall benefits and risks in target populations [8–10]. This assessment typically consists of a series of steps from preclinical studies to the clinical stage. The final step prior to approval decision is a phase III randomized controlled trial (RCT) whereby the candidate is administered to a large group of volunteers, the treatment group, while another group, the control group, receives a placebo or a comparator vaccine. The candidate’s efficacy is assessed by comparing disease incidence in the treatment group with disease incidence in the control group [11]. Vaccine safety is monitored by comparing the incidence of adverse events of interest in the treatment group and in the general population [12, 13]. While ever more advanced methods are being developed for the design and result analysis of RCTs [14], simple efficacy and safety estimands are still used overwhelmingly (see for instance published FDA approval data [15]).

In this article, we use numerical simulations to show with an example that a phase III-like clinical trial may only poorly inform approval and public health decisions in a Disease X outbreak scenario. We model the spread of the disease with a susceptibleinfectious-recovered (SIR) model – a minimal model structure widely used in textbooks to illustrate epidemiological concepts – with parameter uncertainty represented by probability distributions. In our simulated scenarios, a vaccine approval decision is made based on efficacy and safety hypothesis testing in a placebo controlled clinical trial. If the vaccine is approved, it is then delivered to the whole susceptible population. This scenario is simulated for a number of randomly drawn uncertain parameter values, so uncertainty is propagated from, say, biological parameters of Disease X down to the public health impact of vaccine approval decision. The objective is to minimize the total cost of the outbreak, which includes infection costs, vaccination costs, and vaccine adverse events costs. Notice that we take a payer’s or societal perspective rather than an individual’s perspective, which is an important distinction to make when interpreting our results.

Our illustrative example relies on a specific and stylized emergency response scenario where large-scale vaccine rollout is an option. Still, the underlying reasoning applies to any instance of binary (go/no-go) decision under uncertainty during vaccine development. Issues similar to those pointed here can be expected e.g. for decisions based on correlates of protection [16] or non-clinical endpoints (biomarkers) that could be considered in an emergency response scenario. Our scenario is optimistic in this respect, as we assume that a clinical trial is feasible under favorable conditions, which may of course not always be the case. Under this assumption, we elaborate on an example where clinical trial efficacy and safety estimates are actually misleading on average over uncertain parameters. That is, they do not provide “minimum information” [17] to be improved upon, e.g. through post-approval surveillance or the collection of real-world evidence (RWE, see e.g. [18]), but may rather lead to poor and potentially irreversible public health decisions. The claim that clinical trials are imperfect yet better than nothing does not hold in our scenario. Similarly, arguments in favor of efficacy as “an intrinsic property of the vaccine (…) that can be generalized from one setting to another” [19] fall, as our results suggest potential poor performance of approval decisions even assuming that RCT and rollout are carried out in the same setting (but with dynamic effects in the way). We argue that the issue is not specific to our particular example of clinical trial data (mis)use, and goes beyond incremental methodological or technical fixing.

The design and use of clinical trials to support decision making has previously been addressed extensively and across therapeutic areas. General articles (for instance [20] or more recently [21]) provide lists of design mistakes to be avoided, warnings against RCT data misinterpretation, or RCT limitations to overcome. These issues can be solved technically, and the articles’ focus lies accordingly on the technical difficulty of properly estimating efficacy and safety. This misses the point of the actual value of efficacy and safety estimates to support decision making. As for more specific quantitative studies, they are most often solution-oriented and focus each on a limited number of design or analysis issues in specific scenarios. By contrast, we use a quantitative minimal example to make a general point about the value and use of RCT data for decision making. We control merely technical issues by assuming favorable trial conditions, which allows us to dive into more fundamental problems.

Decision making, in our scenario, is undermined by a variety of interrelated factors. One conspicuous barrier to using clinical trial data for proper decision making is that the eventual net benefit of a vaccination campaign depends on parameters that may not be easily (and are typically not) assessed in a clinical trial. This is usually the case of economic parameters in a broad sense (e.g. logistical costs or constraints, averted loss of productivity), but also societal benefits [17, 22, 23]. Collecting economic data alongside otherwise typical clinical trials (“piggybacking”) has long been discussed [24–28] but is not yet widespread [29]. At any rate, it is reasonable to assume remaining uncertainties about economic parameters at the time of approval decision in a Disease X outbreak scenario.

This uncertainty is compounded by uncertainties regarding parameters that *do* influence clinical trial results, for example pathogen, vaccine, or population characteristics, including the potential for indirect vaccine protection, which is typically ignored in vaccine trials [17, 22, 30]. The total net benefit of a vaccination campaign depends on the long-term trajectories of the epidemic with and without vaccination (that is, the difference of the two), that are influenced by these uncertain parameters. Yet parameter values yielding very different dynamics in the long run may result in similar clinical trial results. This issue can be more serious in a Disease X outbreak scenario, because the early stages of an epidemic do not reflect plainly its long-term dynamics in general [31–33].

We emphasize the distinction between two sources of uncertainty that we both tackle with Monte Carlo simulations in our study: first, parameter uncertainty, that is the fact that the true value of the parameters is not known, and, second, uncertainty due to trial random sampling e.g. of a heterogeneous population or of a stochastic process in an otherwise homogeneous population.^1^ Traditional clinical trial design methods have mostly ignored the former and focused on the latter (despite recommendations, see e.g. [26]). Take for example textbook sample size calculations: given a working significance level, and assuming a treatment efficacy value (e.g. the smallest worthwhile value, which is arguably an uncertain parameter at the time of the trial), a sample size is picked to achieve a target power – the limitations of this “conditional power” approach have been discussed by other authors, e.g. [31, 34].^2^ The external validity of trials has largely been equated with proper sampling (inclusion and sample size) of a possibly heterogeneous population e.g. in scholarly discussions about *efficacy* (performance under trial conditions) versus *effectiveness* (performance in the field) of treatments [20]. A large portion of the clinical trial design literature focuses on tweaking existing methods so that sample efficacy and safety estimates accurately reflect efficacy and safety in the real world (that is, improve sampling to avoid biases). However, the actual value of more accurate efficacy and safety estimates to inform decisions is rarely considered in practice. To be fair, an important body of earlier studies used *value of information* reasoning to determine optimal sample sizes considering the cost and benefit of sampling, but in static settings and using analytical methods that would be inappropriate in an infectious disease scenario [43–52] – we refer to [33] for an illustration of the complex dynamics of value of information in a Disease X emergence scenario. In the present study, we go beyond the usual efficacy versus effectiveness distinction and the corresponding problem of appropriate sampling. Specifically, we assume a large sample size in our base case, and minimal population heterogeneity. We make the optimistic assumption that the sampled and general populations have the exact same characteristics, save for vaccination date, and that the only inter-individual differences are differences in stochastic trajectories (infection, recovery) and treatment (vaccination). However, all model parameters are uncertain.

The introduction of Bayesian methods in clinical trials has been a significant step towards proper consideration of parameter uncertainties. Bayesian methods offer ways to aggregate information as it accumulates during a trial [53, 54]. Prior distributions over uncertain parameters are updated into posterior distributions that can in turn be sampled to extrapolate from available information and support decision making e.g. through costeffectiveness analyses [55, 56]. More generally, Bayesian methods “move from the 0–1 utilities to utilities that put values on units of the outcome(s) of interest” (Stangl [57] in her discussion of the seminal paper by Brophy [58]). Implicit 0–1 utilities are found in frequentist hypothesis testing where the hypothesis of treatment effect is accepted or rejected depending on whether the observed difference between the treatment and control groups lies on one side or the other of a predefined threshold. Examples of threshold-based criteria include requirements for COVID-19 vaccine issued in 2020 [59–61]. Thresholds are known to lose information [49, 62, 63]. This loss of information is twofold: first, hypotheses concern only a limited number of summary estimands, while the decision problem involves many parameters as discussed above^3^ and second, information is lost about the level of parameter uncertainty.

Still, Bayesian methods are not widely used in clinical trials [54], and not always to their full potential. For example, Bayesian clinical trials often consist in producing the posterior distribution of a single summary estimand (e.g. efficacy), from which extrapolation can be difficult – see [65] for instance. Similarly, the value of information studies cited above [43–52] did build upon Bayesian reasoning, but with a limited number of uncertain parameters and a limited propagation of uncertainty. As for implications, they mostly criticized thresholds used in hypothesis testing for being arbitrary; we go a bit further and show that they may also be misleading. Finally, it is worth mentioning the substantial Bayesian literature on predictive probabilities of trial success (including frequentist trials) to support interim decision making [34, 53, 66, 67]. Interestingly, some authors in this literature propagated biological uncertainties down to trial performance and considered a larger number of parameters, but not economic parameters [68, 69]. While we take a public health perspective in this article, some of our comments may still apply to trial sponsor decision making.

Our article is closer to some previous studies. Hill-McManus and Hughes [63] used RCT simulations to pick a trial design (inclusion criterion, sample size, and medicine doses) maximizing the manufacturer’s return on investment. They propagated biological uncertainties down to the marketing stage, and modeled reimbursement decisions based on a cost-effectiveness model. While they considered a wide range of uncertain parameters and did include economic parameters, those are not uncertain in their study. They also assumed seamless integration of RCT results with health economic modeling and reimbursement decision, which may not be the rule in practice. To some extent, our work is in line with Perkins et al. [70] (unpublished to our knowledge). This study argued that “statistical uncertainties” (i.e. around efficacy estimates) and “biological uncertainties” (i.e. about the disease and vaccine mode of action) make it difficult to make public health decisions based on the results of a dengue vaccine clinical trial alone. Our methods differ, however, as they back-calculated uncertain parameter distributions based on the results of a real clinical trial. By contrast, we start from hypothetical uncertain parameter distributions and simulate RCTs *as if* we were faced with Disease X outbreaks. They also focused on vaccine efficacy and ignored safety while we consider both, and did not include economic parameters while we do.

The remaining sections of the article are as follow. Section 2 introduces all modeling assumptions and modeled scenarios. The results are presented in Section 3. Section 3.1 shows how making a decision based on RCT results compares to other policies in our scenario. In Section 3.2, we vary the design of RCTs – Section 3.2.1 is the core of the article and looks into statistical tests, while Section 3.2.2 describes more concisely the influence of trial duration and sample size. Section 4 concludes.

## 2 Materials and methods

### 2.1 Disease X transmission model

We consider the emergence of an unknown infectious disease, Disease X, in a closed homogeneous population of *N* = 10^8^ individuals. We assume that the spread of the disease can be described with a deterministic SIR (susceptible-infectious-recovered) compartmental model with basic reproduction model *R*_0_ and 1*/γ* average duration of infectious period. Figure App-1 shows a sketch of the transmission model.

At date 0, a single individual is infected and all other individuals are susceptible. A vaccine is available from date *T*_*v*_ = 900 days (unless otherwise specified). We assume no alternative vaccine or treatment. The vaccine is administered as a single dose and protects vaccinated individuals with probability *p*. Vaccine protection is immediate and there is no waning of protection. Adverse events occur in vaccinated individuals with probability *ϵ*, in which case the average duration between vaccination and adverse events is *δ*. Adverse event probability *ϵ* is independent of vaccination success.

The cost of infection is *c*_*i*_ per infectious day, and the cost of adverse events is *c*_*ae*_ per case. The total cost of producing and administering one vaccine dose is *c*_*v*_ – we ignore fixed costs for concision, which amounts to assuming that they have already been incurred at time *T*_*v*_.

### 2.2 Decision problem

At time *T*_*v*_, a decision must be made by the decision maker (health authorities or the payer) between several options. They can implement vaccination for the entire susceptible population immediately (emergency vaccination); they can decide to not vaccinate anyone; or they can implement a RCT first and implement vaccination later conditionally on the RCT outcomes (by ways to be defined below). We assume that the objective of the decision maker is to minimize the expected total cost over the course of the epidemic.

### 2.3 Uncertainties and prior information

We consider parameter uncertainties at time *T*_*v*_ represented by the parameter prior distributions shown in Table 1. The population size *N* and the date *T*_*v*_ from which the vaccine is available are assumed to be known. We are only interested in decision making from date *T*_*v*_. The value of *T*_*v*_ clearly has an influence on the trade-off between emergency vaccination and clinical research, but this was investigated elsewhere [33] and is left outside the scope of the present study. Figure 1 shows 400,000 parameter draws from the prior distributions of uncertain parameters. We will simulate decision making for each parameter draw.

**Table 1.** Model parameters prior distributions. Time unit: day. Cost unit: 10^3^ USD.

| Parameter | Distribution | Truncation |
| --- | --- | --- |
| $N$ | $10^8$ (fixed) | |
| $T_v$ | 900 (fixed) | |
| $R_0$ | Gamma distribution with mean 1.05 and standard deviation 0.2 | $[1.01; \infty]$ |
| $\gamma$ | Lognormal distribution with mean 0.1 and standard deviation 0.1 | $[0.02; \infty]$ |
| $p$ | Uniform distribution | $[0, 1]$ |
| $\epsilon$ | Lognormal distribution with mean 0.003 and standard deviation 0.003 | $[0; 1]$ |
| $\delta$ | Normal distribution with mean 40 and standard deviation 20 | $[0; \infty]$ |
| $c_v$ | Normal distribution with mean 0.1 and standard deviation 0.1 | $[0; \infty]$ |
| $c_i$ | Normal distribution with mean 0.1 and standard deviation 0.03 | $[0; \infty]$ |
| $c_{ae}$ | Normal distribution with mean 50 and standard deviation 50 | $[0; \infty]$ |

**Figure 1.**
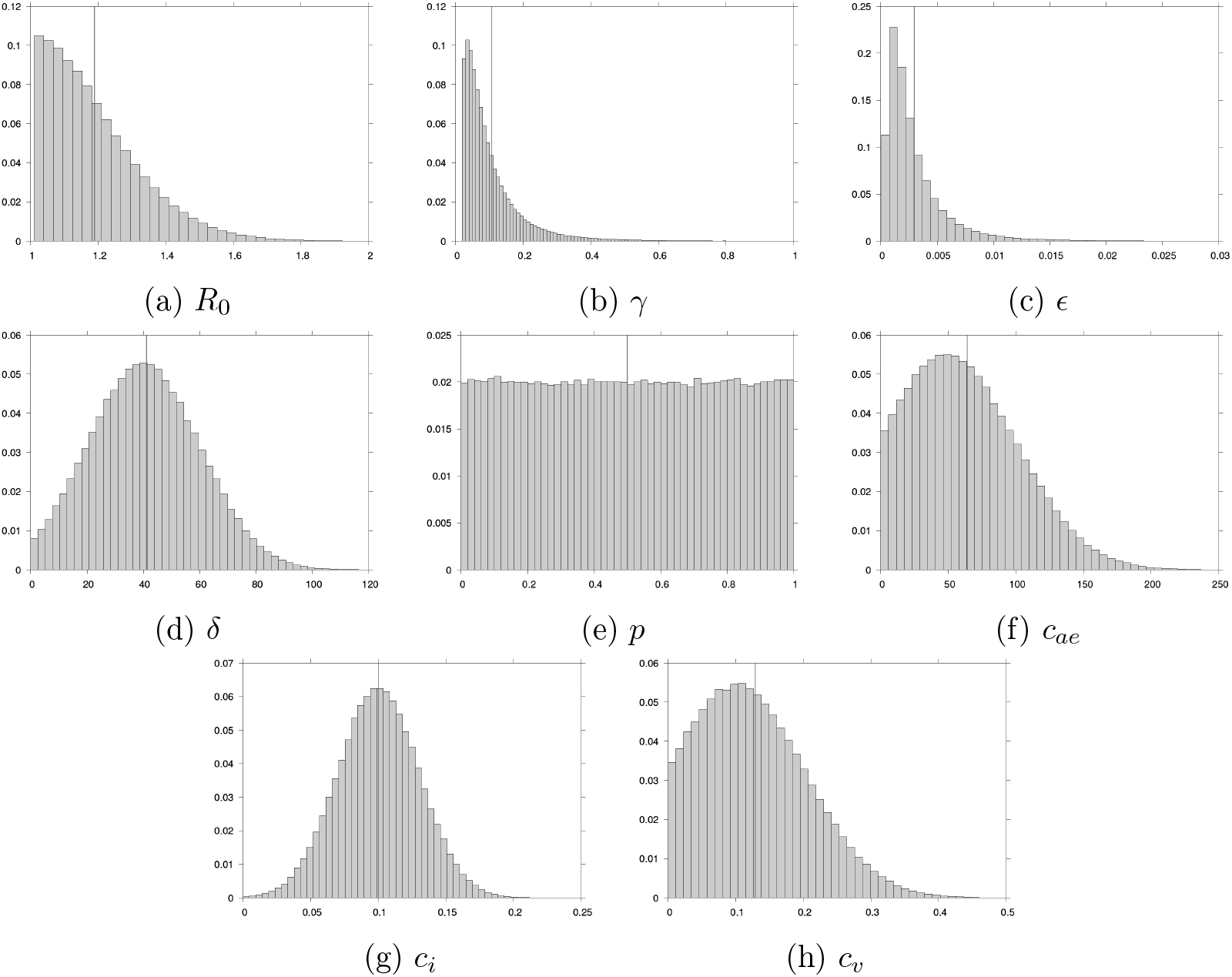
Frequency distributions for 400,000 random draws of parameter values. Vertical lines show average values. Time unit: day. Cost unit: 10^3^ USD.

Prior information includes knowledge of the model structure, that is SIR transmission in the absence of intervention, and knowledge of the vaccine characteristics (all-or-nothing protection, no waning). These assumptions can readily be relaxed by extending the model with additional compartments and uncertain transition rates, or by introducing several models with a probabilistic weight for each. In this didactic article, the SIR structure is to be understood as a minimal working model of transmission in a population.

We choose biologically plausible disease parameter values (*R*_0_, *γ*). The values of *ϵ* and *δ* are assumed, while the prior distribution of *p* represents minimal prior information on vaccine efficacy. The order of magnitude of *ϵ* may seem high (see [71, 72]), but recall that it represents the probability of a “synthetic” adverse event that we hypothesized for concision, whose probability is the sum of several “real” adverse events of interest. The order of magnitude of the cost *c*_*ae*_ of adverse events is based on damage payments in the US and the UK, with lower range values representing mild uncompensated adverse events [73, 74]. The order of magnitude of the cost *c*_*i*_ of an infectious day is that of average daily wages in high-income countries [75]. The cost per administered vaccine dose *c*_*v*_ derives from vaccine purchase prices [76, 77]. We emphasize that prior distributions represent available information or beliefs *before* vaccine approval decision. In addition, this information concerns one particular outbreak and one particular vaccine candidate. For these two reasons, prior distributions may not correspond to distributions observed among approved vaccines or inferred from past epidemiological data.

### 2.4 Simulated RCTs and trial simulation method

We simulate two-arm clinical trials starting at date *T*_*v*_ with duration *T*_*ct*_. The default trial duration is *T*_*ct*_ = 80 days. The sample size is *N*_*ct*_ (default *N*_*ct*_ = 50,000), and the treatment and control arms have equal size *N*_*ct*_*/*2. Individuals in the treatment arm receive the vaccine, while individuals in the control arm receive a placebo.

The number of infections in the treatment and control arms, 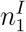 and 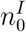 respectively, and the number of adverse events *n*_*ae*_ in the treatment arm are tracked for the duration of the trial *T*_*ct*_. In trial simulations, infections are randomly drawn at each time step in a binomial distribution with probability *R*_0_*γI/N* among the remaining susceptible individuals in the considered arm. The number *n*_*ae*_ of adverse events in the treatment arm is drawn in a binomial distribution with parameters *N*_*ct*_*/*2 and 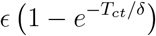.

Vaccine efficacy and safety are then tested at time *T*_*v*_ + *T*_*ct*_. We test whether vaccine efficacy is *superior* [78] to that of the placebo using a z-test, where the null hypothesis 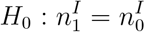 is tested against hypothesis 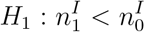 with significance level *α*. The rate of occurrence of adverse events is estimated as 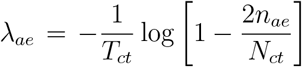. The vaccine passes the safety test if *λ*_*ae*_ is below a threshold 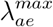 based on the background incidence of medical conditions of interest in the unvaccinated population. See [12, 71, 72, 79, 80] for illustrations and discussions of this approach.

In the following we will focus on two particular clinical trials, denoted CT95 and CT0. In both trials, the sample size and duration are set to their default values *N*_*ct*_ = 50,000 and *T*_*ct*_ = 80 days. The safety threshold is set to 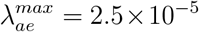 day^-1^.^4^ CT95 and CT0 differ in the significance level used in efficacy testing. For CT95, significance is set to customary value *α* = 0.05 (95% confidence), while for CT0 it is set to *α* = 1 (0% confidence). With CT0, all vaccines pass the efficacy test and may only be rejected on the basis of safety.

## 3 Results

### 3.1 Use of RCT information

The objective of a RCT is to evaluate the safety and efficacy of a vaccine candidate. Table 2 shows how this information can help reduce the total cost over the course of an epidemic. We estimate the expected total costs over prior parameter distributions in different intervention scenarios (Figure App-2 shows the full cumulative distributions of total costs). The simulated intervention scenarios are as follow. In scenario “no trial, no vacc.” no one is vaccinated. “No trial, vacc. at *T*_*v*_” corresponds to emergency vaccination: all susceptible individuals are vaccinated at time *T*_*v*_ i.e. as soon as the vaccine is available. Under policies “CT0” and “CT95”, a clinical trial is implemented at time *T*_*v*_. If the vaccine passes the safety and efficacy tests, as defined in Section 2.4, all susceptible individuals receive the vaccine at time *T*_*v*_ + *T*_*ct*_.

**Table 2.**
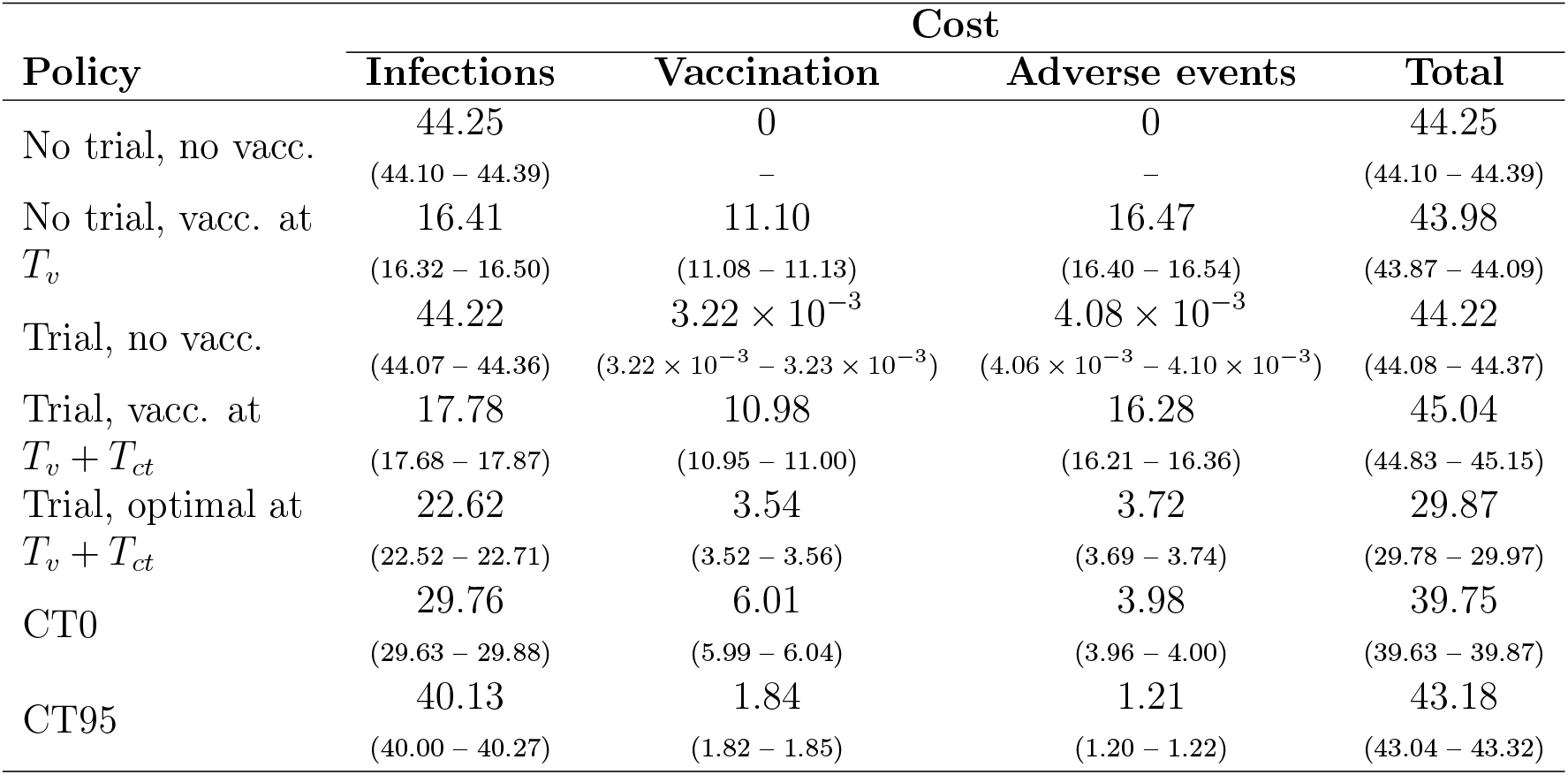
Summary simulation results for different vaccination policies. Costs in 10^9^ USD. Averages over 400,000 parameter draws, and one simulation per parameter draw. Parentheses: 95% confidence intervals.

In scenarios “trial, no vacc.” and “trial, vacc at *T*_*v*_ + *T*_*ct*_”, a trial is implemented at time *T*_*v*_ but the vaccine is used or not irrespective of the trial result. Comparing “no trial, no vacc.” and “trial, no vacc.” shows that the influence of the clinical trial (that is, of vaccinating *N*_*ct*_*/*2 = 25,000 individuals at time *T*_*v*_) on disease transmission is small in our scenario. Similarly, comparing emergency vaccination (“no trial, vacc. at *T*_*v*_”) to “trial, vacc. at *T*_*v*_ + *T*_*ct*_” shows that delaying vaccination by *T*_*ct*_ = 80 days to implement a RCT has little influence on the course of the epidemic.

Making a decision based on CT0 or CT95 allows us to reduce the overall cost compared to making a decision irrespective of trial results. Compared to “trial, no vacc.”, the increase in vaccination and adverse event costs is more than compensated by the decrease in infection costs. Compared to “trial, vacc. at *T*_*v*_ + *T*_*ct*_” and to emergency vaccination, the increase in infection costs is more than compensated by the decrease in vaccination and adverse event costs.

“Trial, optimal at *T*_*v*_ + *T*_*ct*_” is a hypothetical scenario in which a clinical trial is implemented at time *T*_*v*_, and the true value of all uncertain parameters is known at time *T*_*v*_ +*T*_*ct*_, so that the best policy can be picked between vaccinating and not vaccinating the remaining susceptible individuals. This gives a (loose) lower bound on the total cost achievable through clinical research. Using the no intervention scenario (“no trial, no vacc.”) as a baseline, the value brought by CT0 and CT95 is worth about 31 % and 7 % respectively of the value of perfect information.

Noticeably, we obtain a better result with CT0 compared to CT95. Recall that CT0 only tests safety, while CT95 also tests efficacy with significance level *α* = 0.05. A decision maker would arguably be more likely to implement the customary CT95 trial rather than CT0 *a priori*, that is without performing the analyses presented in Table 2. It is critical to fully consider uncertainties in these analyses. Table App-1 is the equivalent of Table 2 but with uncertain parameters set to their prior expected value. On this basis, the best policy would be to do nothing.

CT0 and CT95 are compared in more details in Table 3. In our scenario, vaccination is optimal at time *T*_*v*_ + *T*_*ct*_ for 31.80 % of uncertain parameter values, but CT0 and CT95 poorly prescribe vaccination when it is actually optimal. The true positive rate of CT95, in particular, is only 6.16*/*31.80 ≈ 0.19, and that of CT0 is 20.47*/*31.80 ≈ 0.64. Because CT95 is more restrictive, it has a lower false positive rate of 10.86*/*68.20 ≈ 0.16 compared to CT0 (34.25*/*68.20 ≈ 0.50).

**Table 3.** Correlation matrix between vaccination policy implemented based on CT0 and CT95 (row) and vaccination policy implemented under perfect information after a clinical trial (column). Percentages of 400,000 parameter draws.

|  |  | Vaccination | No vaccination | Total |
| --- | --- | --- | --- | --- |
| CT0 | Vaccination | 20.47 % | 34.25 % | 54.72 % |
|  | No vaccination | 11.33 % | 33.96 % | 45.29 % |
|  | Total | 31.80 % | 68.20 % | 100.00 % |
| CT95 | Vaccination | 6.16 % | 10.86 % | 17.02 % |
|  | No vaccination | 25.63 % | 57.34 % | 82.97 % |
|  | Total | 31.80 % | 68.20 % | 100.00 % |

### 3.2 Clinical trial design

The performance of a clinical trial depends on its design. In this section we ask whether the performance of CT0 and CT95 could be improved on average over uncertain parameters by changing the statistical tests, the sample size, or the trial duration.

#### 3.2.1 Statistical tests

Here, we first describe how safety and efficacy threshold values used in statistical tests may increase or decrease the expected total cost of making a decision on the basis of a RCT, which will tell us about RCT design optimization on average over uncertain parameters. This is shown in Figures 2–4. Then, we describe the relationship between safety and efficacy estimates (i.e. RCT observations), and the total benefit of vaccinating the whole population, *given* uncertain parameter values. This will help understand the previous results and give insights into the level of information provided by a RCT in our scenario. This is done in Figures 5 and 6.

**Figure 2.**
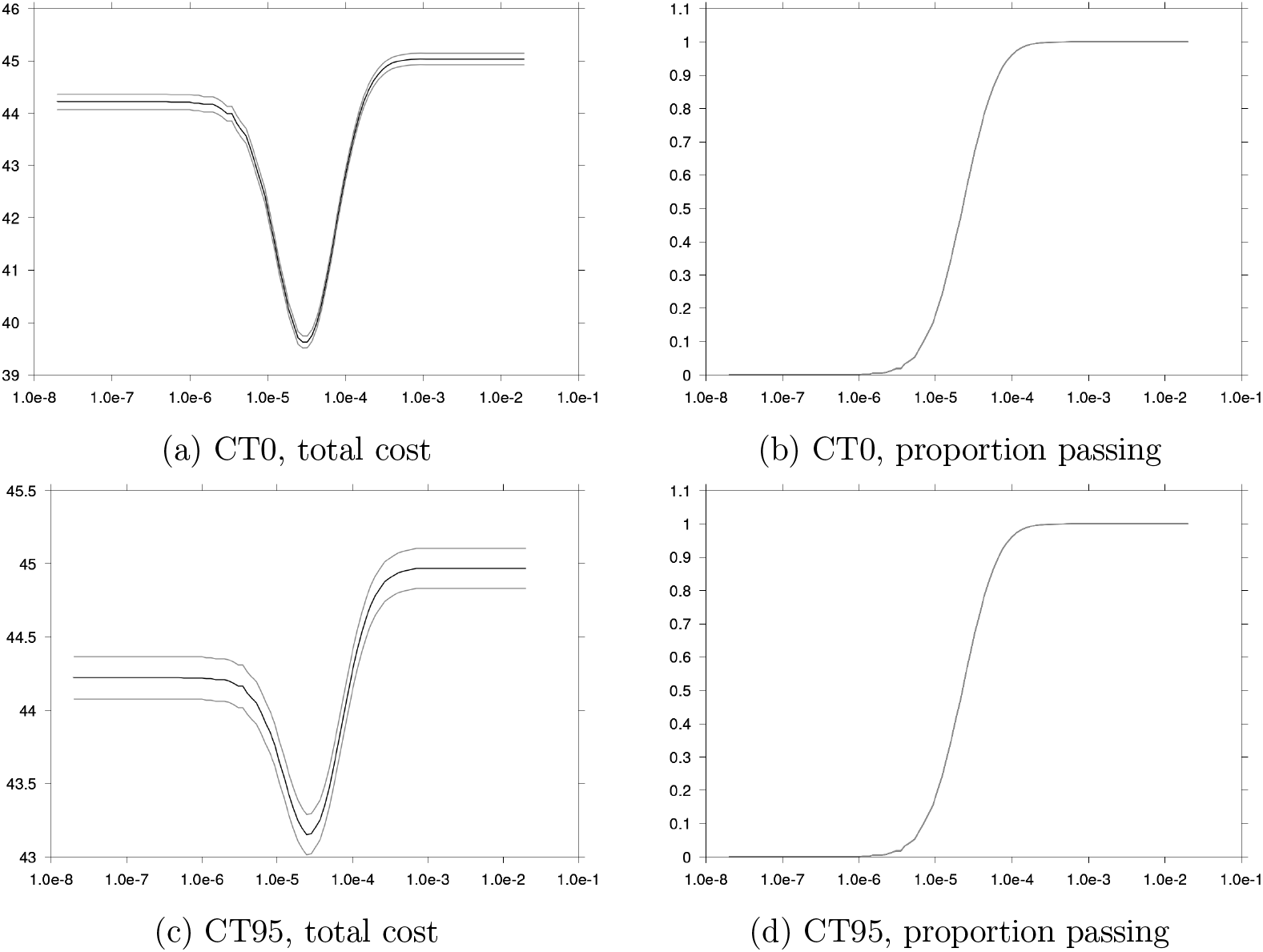
Expected total cost (left panels, 95% CI in light lines, cost in 10^9^ USD) and proportion of simulations passing the safety test (right panels) as functions of safety threshold 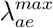 (log_10_ scale).

**Figure 3.**
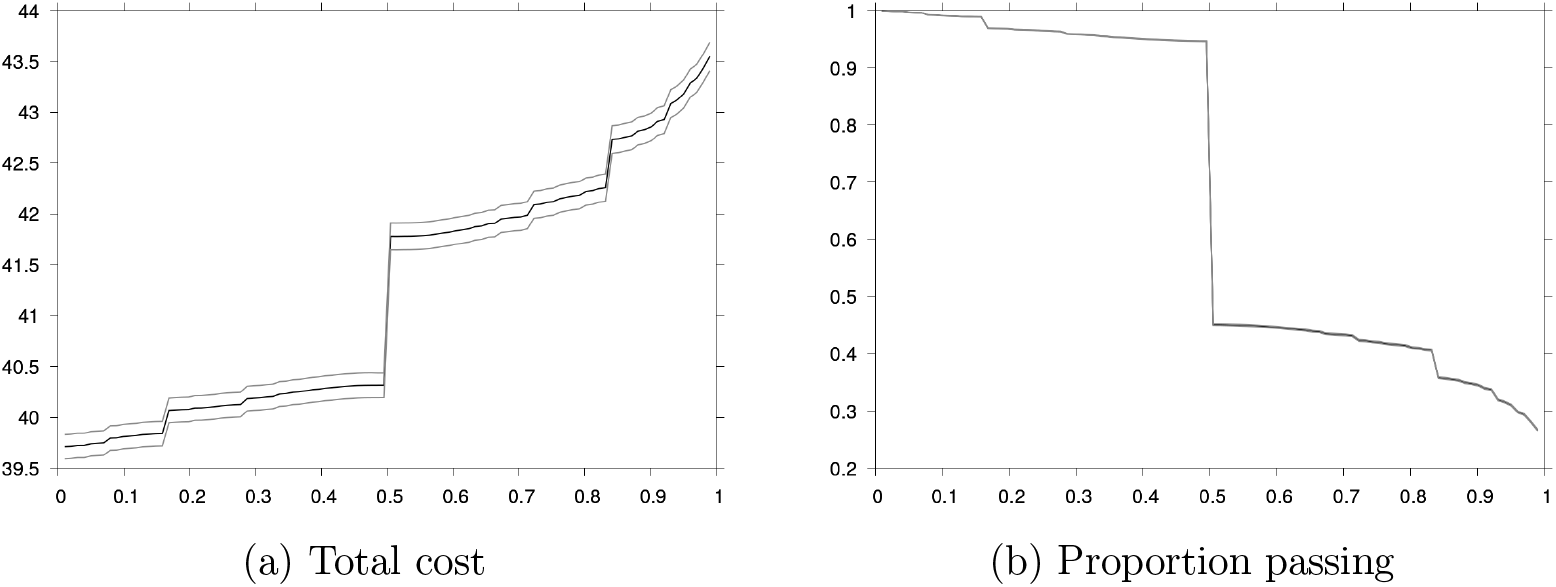
Expected total cost (left, 95% CI in light lines, cost in 10^9^ USD) and proportions of simulations passing the efficacy test (right) as functions of the efficacy test confidence level 1 − *α*.

**Figure 4.**
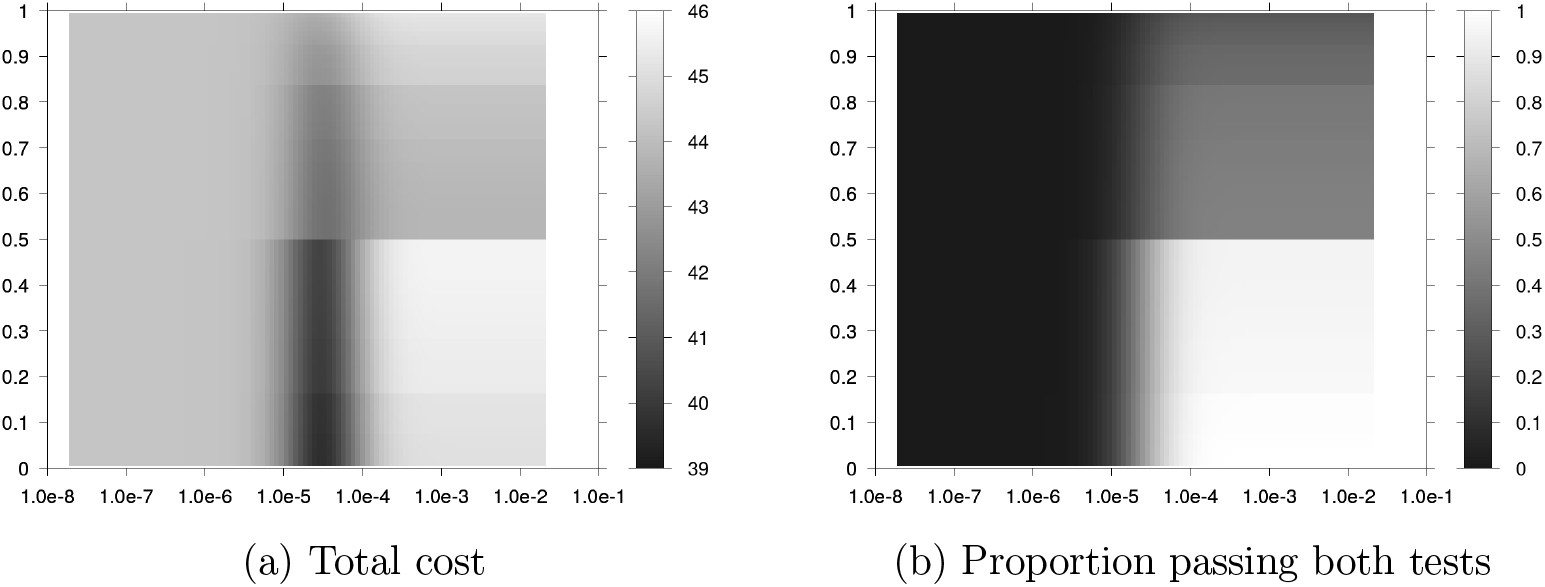
Expected total cost (left) and proportion of simulations passing both safety and efficacy tests (right) as functions of safety threshold 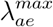 (x-axis, log_10_ scale) and efficacy test confidence level 1 − *α* (y-axis).

**Figure 5.**
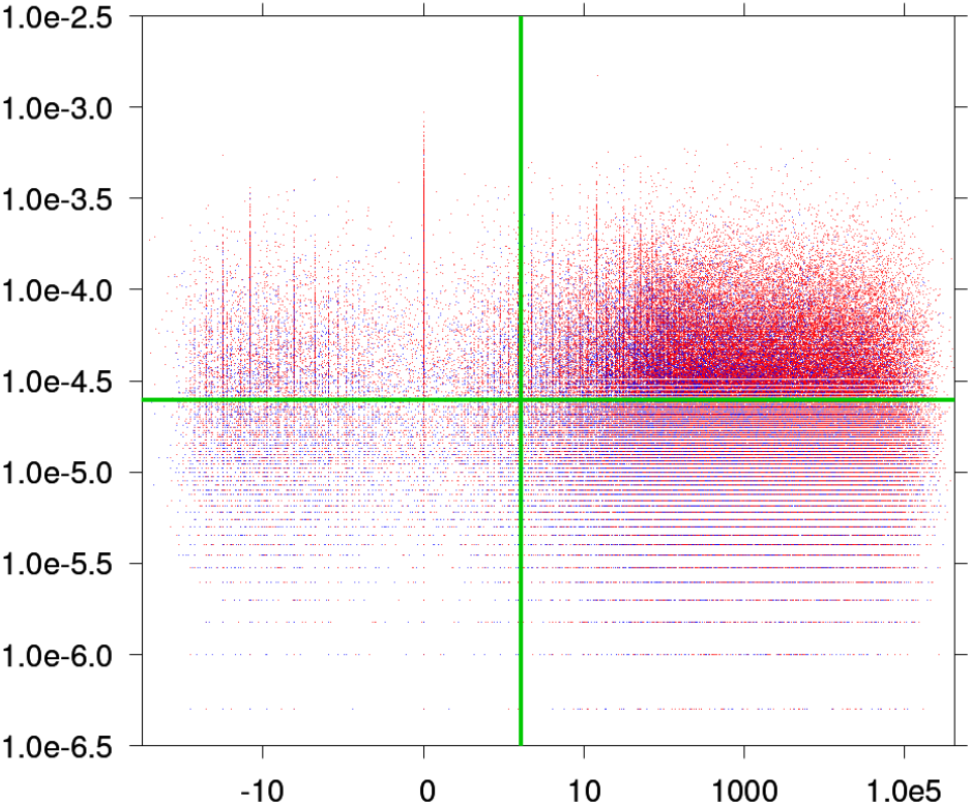
Optimal policy between “trial, no vaccination” (red) and “trial, vaccination at *T*_*v*_ + *T*_*ct*_” (blue) as a function of the measured adverse event rate *λ*_*ae*_ (y-axis, log_10_ scale) and the efficacy 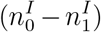 z-value (x-axis, symlog scale). 400,000 parameter draws and one trial simulation per draw. Green lines: safety and efficacy thresholds of CT95.

**Figure 6.**
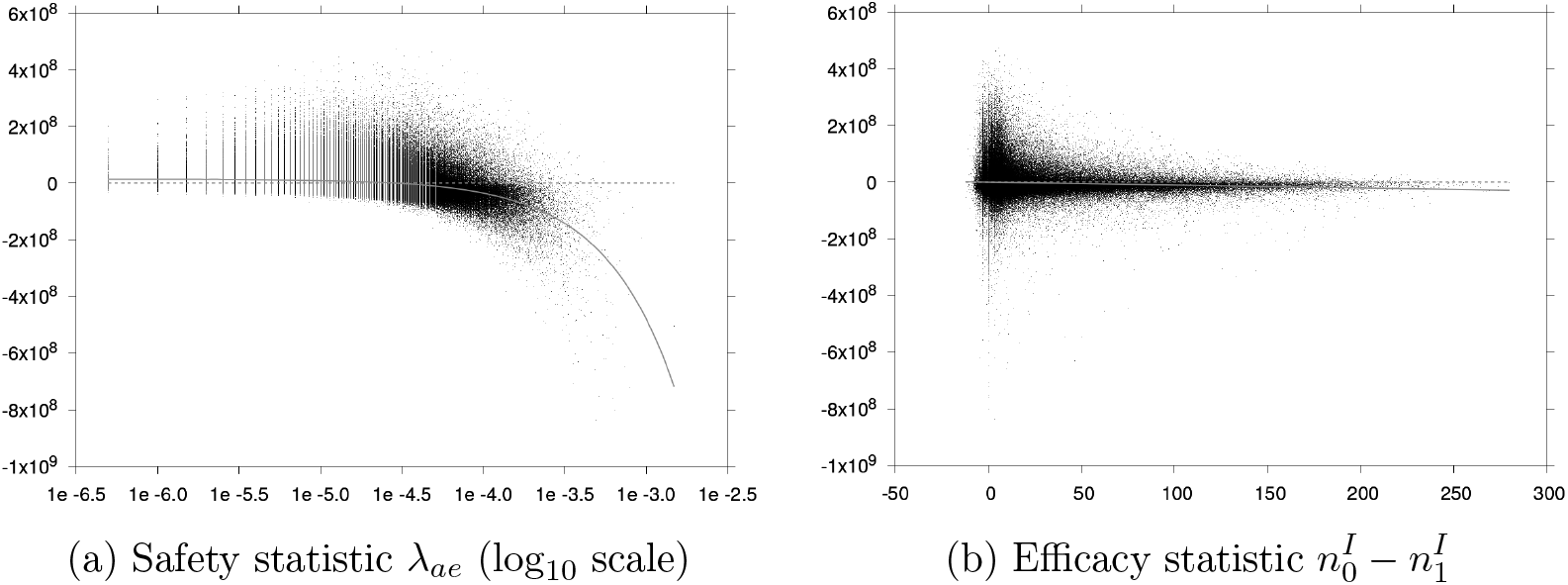
Difference in total cost over the epidemic between policies “trial, no vaccination” and “trial, vaccination at *T*_*v*_ + *T*_*ct*_” as a function of the measured adverse event rate *λ*_*ae*_ (left) and the difference in numbers of infections between the control and treatment arm 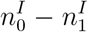 (right). 400,000 parameter draws and one trial simulation per draw. Grey line: best fit linear regression. Dashed line: *y* = 0 line.

Let us look into the influence of safety threshold 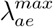 on clinical trial performance. In Figure 2, we show for both CT0 and CT95 the expected total cost and the proportion of simulations (one simulation for each of the 400,000 parameter draws) in which the vaccine passes the safety test as a function of 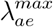. If the threshold is too low, the vaccine is rejected with probability 1 and we obtain the same total cost as with policy “trial, no vaccination” in Table 2. If the threshold is too high, the vaccine is rejected with probability 0 on the basis of safety (with CT95, however, it may be rejected on the basis of efficacy): with policy “CT0”, we obtain the same cost as with policy “trial, vaccination at *T*_*v*_ + *T*_*ct*_”. The threshold is optimal for an intermediate value.

Let us now turn to the influence of the confidence level 1 − *α* used for efficacy testing. Figure 3 shows the expected total cost and probability of passing the efficacy test as functions of 1 − *α* for our baseline 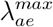 value. Recall that 1 − *α* is equal to 0 and 0.95 for CT0 and CT95 respectively. In our scenario, the more restrictive the efficacy test, the higher the overall cost. The curves are not smooth because several simulations may have the exact same difference in number of infections between the treatment and control arms. In our scenario, this is obtained for small numbers of infections in both arms. For example, there is a single infection case in the control arm and none in the treated arm for more than 4% of simulations, and we obtain the opposite result for more than 2% of simulations. The jump at 1 − *α* = 0.5 is explained by a large number of simulations with the same number of cases in both arms. In particular, there is no infection case in either arm for about 32% of the simulations.

Figure 4 shows how varying both 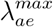 and 1 − *α* affects the results – see also receiver operating characteristic (ROC) curves in Section C in Appendix. The higher expected total cost is incurred for too permissive trials (bottom right quadrant). Starting from there, it is better to optimize 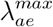 *alone* rather than increasing stringency of both efficacy and safety testing. For permissive safety tests (right part of the heat map), the trial can be improved by making the efficacy test more restrictive (top right quadrant). However, for intermediate (and optimal) safety threshold values, the more restrictive the efficacy test, the higher the overall cost.

In fact, threshold-based efficacy testing is not a good tool to inform public health policies. Let us dive into how RCT (observed) safety and efficacy estimates relate to the actual optimal policy between vaccinating and not vaccinating after the trial. For 400,000 parameter draws and trial simulations we computed (i) the optimal policy between “trial, no vaccination” and “trial, vaccination at *T*_*v*_ + *T*_*ct*_”, and (ii) the trial test statistics used for safety and efficacy testing, *λ*_*ae*_ and 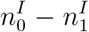 (and the efficacy z-value, to be specific) respectively. Figure 5 shows which of the two policies is optimal for each simulation in the safety-efficacy plane. The test thresholds of CT95 are shown in green. In the bottom right quadrant scenarios, the vaccine would be approved on the basis of CT95. In the two top quadrants, the vaccine fails the safety test. In the two left quadrants, the vaccine fails the efficacy test. Red dots in the bottom right quadrant correspond to false negatives (the vaccine is rejected while it should be approved). Blue dots in the other quadrants correspond to false positives (the vaccine is approved while it should be rejected). We emphasize that each dot corresponds to a random draw of uncertain parameters and trial sample. In particular, the actual RCT outcomes would correspond to a single dot. Arguably, no pair of threshold values can adequately separate red dots from blue dots in the safety-efficacy plane. This illustrates the limitations of low-dimensional methods when solving a complex high-dimensional problem. The issue is fundamental, and beyond tweaking threshold values in statistical tests.

Now, how do RCTs perform on average depending on threshold values? For the same 400,000 simulations, we computed the difference in total cost between the two policies – “trial, no vaccination” and “trial, vaccination at *T*_*v*_ +*T*_*ct*_” –, that is the benefit of vaccinating following the trial. We regressed the benefit of vaccination on test statistics as shown in Figure 6. The regression on *λ*_*ae*_ (Figure 6a) crosses *y* = 0 for a cutoff value and has a negative slope. As expected, this suggests that the higher the measured adverse event rate *λ*_*ae*_, the less beneficial the expected value of vaccination. It also suggests that, all other things being equal, there exists an optimal value above which it is optimal to reject vaccination and below which it is optimal to implement vaccination.

By contrast, as displayed in Figure 6b, the regression of the benefit of vaccinating following the trial on 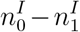, is strictly negative and does not crosse the *y* = 0 line. Hence, on average, vaccinating at time *T*_*v*_ + *T*_*ct*_ is less beneficial in cases where the reduction of infections in the treatment arm compared to the control arm is larger. Hence, as previously shown, the stricter the efficacy test (larger threshold), the worse the average performance of vaccination with vaccines passing the efficacy test (those for which 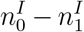 is above the threshold). Also, the regression line not crossing the *y* = 0 line suggests that on average, all other things being equal, it is a better policy not to vaccinate and no threshold policy (even one with vaccination *below* the threshold) can outperform this.

Figure 6 also illustrates the role of parameter uncertainty. As a thought experiment, assume that all parameters are known and the only source of noise is trial participant sampling.^5^ Assume also that the trial has no influence on the spread of the disease. These assumptions are customary in RCT design, particularly when using a conditional power approach for sample size computation. However, under these assumptions, the scatter plots in Figure 6 would reduce to horizontally aligned dots, that is a cross-section of the full scatter plots. Optimizing the statistical power based only on this limited cross-section can obviously lead to suboptimal trial designs under parameter uncertainty.

#### 3.2.2 Trial duration and sample size

In Figure 7, we show how modifying the duration *T*_*ct*_ of the clinical trial or the number *N*_*ct*_ of individuals included in the trial affects the results of policies “CT0” and “CT95” consisting in implementing trials CT0 and CT95 respectively at time *T*_*v*_ and vaccinating at time *T*_*v*_ + *T*_*ct*_ if the vaccine passed the safety and efficacy tests. Recall our baseline values for CT0 and CT95: *T*_*ct*_ = 80 days and *N*_*ct*_ = 50,000 individuals.

**Figure 7.**
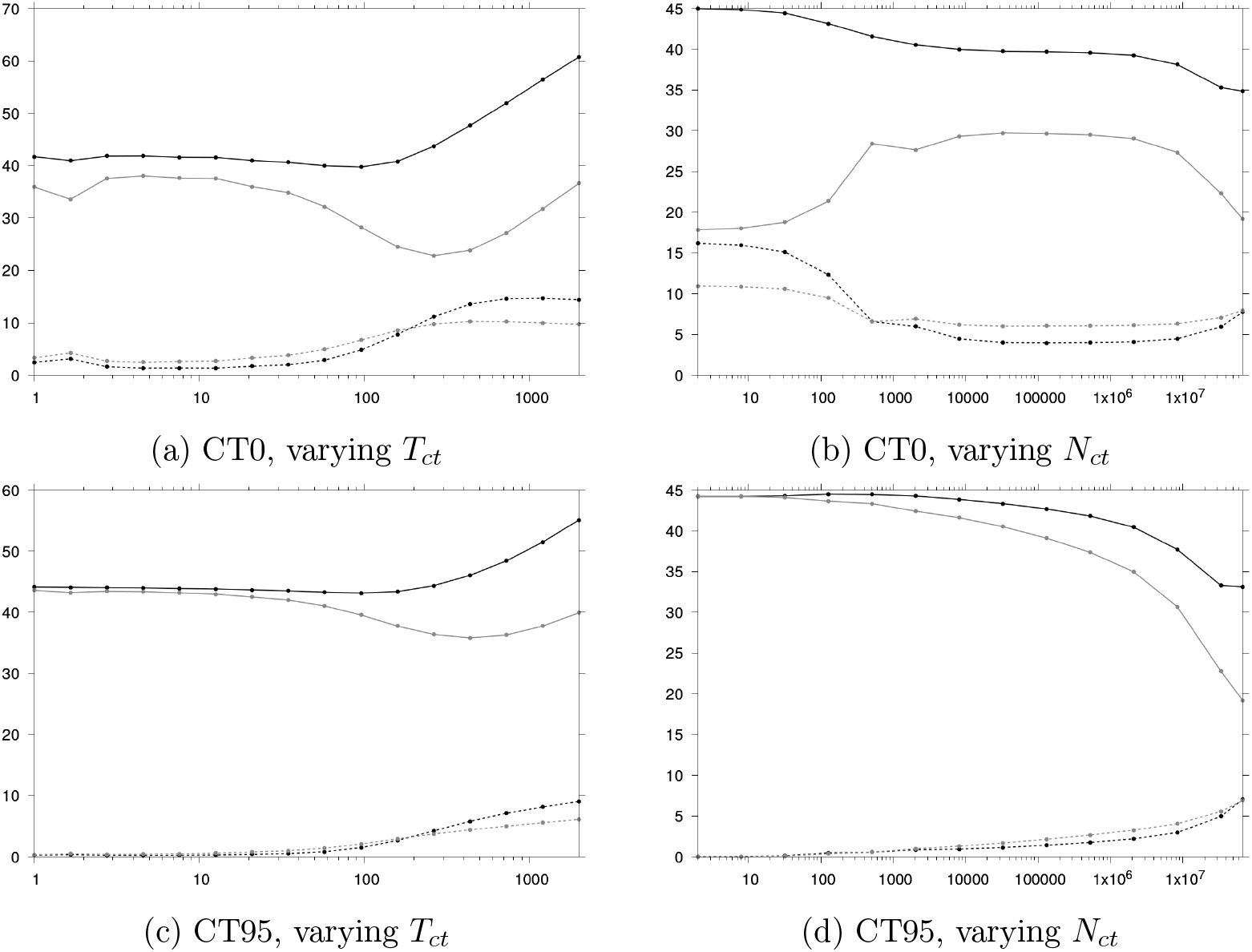
Expected costs (10^9^ USD) as functions of the duration *T*_*ct*_ of the clinical trial (days, left) and the number *N*_*ct*_ of individuals included in the trial (right) for policies “CT0” (top) and “CT95” (bottom). Black: total cost, grey: infection cost, black dashed: adverse event cost, grey dashed: vaccination cost.

Many effects come into play when changing the trial duration and sample size. For instance, both parameters influence naturally the quality of information brought by the trial in terms of efficacy as well as safety. But they also influence the spread of the epidemic and the number of individuals still susceptible and hence who can be vaccinated if the vaccine is approved. These individuals are also those who can still suffer adverse events. The resulting total effect on trial performance is difficult to anticipate *a priori*, which highlights the importance of epidemiological simulations in Disease X clinical trial design.

Also, Figure 7 shows that our baseline *T*_*ct*_ and *N*_*ct*_ values are such that policies “CT0” and “CT95” perform as good as possible in a realistic range.^6^ Hence, the results presented in this article and in particular the poor information given by the RCTs cannot be attributed to poor RCT design.

## 4 Conclusion

We showed with a numerical example that customary vaccine RCTs with hypothesis testing of efficacy and safety may only poorly inform public health decisions in a Disease X outbreak scenario. We modeled the spread of a hypothetical disease under parameter uncertainty, and we simulated various RCTs and vaccine approval policies. Our analysis considers issues (dynamic effects, uncertainties, large number of parameters involved) that are generally discussed separately, either in general terms, or quantitatively but in a limited setting (e.g. with the objective of solving a specific case). We took a public health perspective, with the objective of minimizing the total cost of the epidemic (including the cost of the RCT). We propagated parameter uncertainties e.g. from biological uncertainties down to the total cost of the epidemic. We simulated counterfactuals that never get to be observed in the field – the hypothetical performance of rejected vaccines. This allowed us to properly estimate the expected total cost of the epidemic under different approval scenarios.

Our analysis relies on several optimistic assumptions – that a RCT with a large sample size is feasible, which may not be the case in a crisis, that population heterogeneity is minimal, and that the RCT and vaccine rollout are carried out in similar settings. These assumptions allow us to overlook technical issues that have already largely been tackled elsewhere, and to focus with a minimal example on more fundamental issues that may derail decision making.

We simulated two baseline RCTs: a customary safety and efficacy trial, CT95, and a trial testing safety only, CT0. In our scenario, CT0 enabled better decisions than CT95, on average over uncertain parameters. Compared to doing nothing (no RCT, no vaccination), the reduction in total cost with CT0 and CT95 corresponded to about 31% and 7% respectively of the reduction allowed by hypothetical optimal decisions. Testing safety only was a better option on average than testing both efficacy and safety.

In our scenario, safety testing could be optimized to minimize the expected total cost over uncertain parameters. By contrast, and somewhat counter-intuitively, the more stringent the efficacy test, the higher the expected cost of making a decision based on the trial. We looked into how summary efficacy and safety estimates relate to the actual benefit of vaccinating the population for a sample of randomly drawn parameter values. This highlighted the inadequacy of RCT and threshold-based rules to support decisions in our scenario. Finally, we described the complex interaction of different competing effects when varying the trial duration and sample size.

All results presented in this article depend on modeling assumptions, and on the assumed parameter prior distributions in particular. The article is intended to be didactic and should be understood as a call for the development of more advanced pattern recognition methods in RCTs. Put simply, it illustrates that a complex high-dimensional problem may not easily be solved with simple methods leveraging only two dimensions (efficacy and safety).

We believe that our work may have implications beyond Disease X preparedness and outbreak response. Decision making under uncertainty, in a dynamic setting, and with many parameters involved is ubiquitous at all stages of vaccine development. This is not only true in the case of Disease X response. Uncertainty and dynamic effects may arise from the emergence of variants and seasonality, as is the case for the flu. Finally, workarounds such as non-clinical endpoints that have been proposed for use in emerging disease response may not be silver bullets: while some of the dynamic effects and uncertainty (e.g. whether trial participants will be infected or not) are removed from the problem, some uncertainty (e.g. about how non-clinical outcomes relate to clinical outcomes) is also added. These considerations open exciting avenues for further research.

## Data Availability

All data produced in the present study are available upon reasonable request to the authors

## A Additional figure: transmission model

**Figure App-1.**
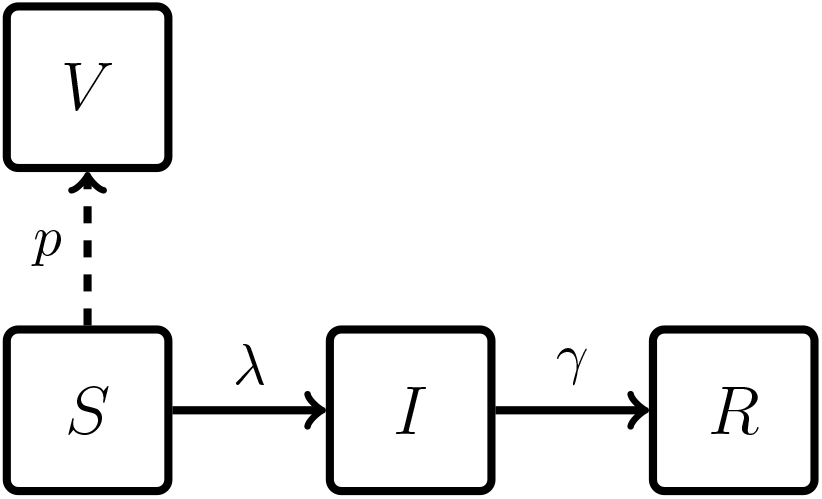
Sketch of the transmission model. Dashed: possible vaccination (instantaneous). 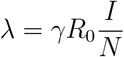 is the force of infection.

## B Additional analyses: baseline scenario

Figure App-2 shows total cost cumulative distributions obtained in our baseline scenario (*N*_*ct*_ = 50,000, *T*_*ct*_ = 80 days). The curve for “trial, vaccination at *T*_*v*_ + *T*_*ct*_” cannot be distinguished from the curve for “no trial, vaccination at *T*_*v*_”. The curve for “no trial, no vaccination” cannot be distinguished from the curve for “trial, no vaccination”.

**Figure App-2.**
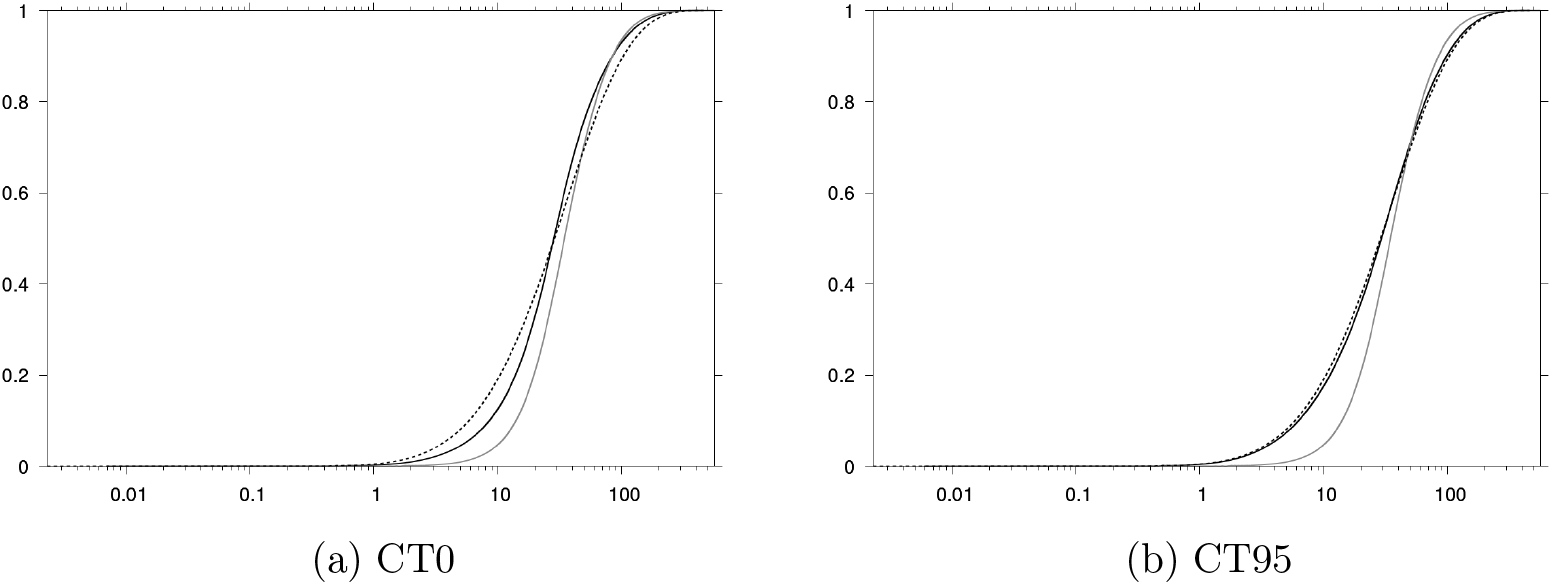
Total cost (10^9^ USD) cumulative distributions for vaccination policies: clinical trials CT0 and CT95 (black), vaccination without trial “no trial, vaccination at *T*_*v*_” (grey) and no vaccination with trial “trial, no vaccination” (dashed black). Curves obtained from 400,000 parameter draws.

Table App-1 shows estimated cost assuming that all uncertain parameters (including efficacy and adverse event parameters) are equal to their prior expected value. Under this assumption, the best policy is to do nothing. Implementing a clinical trial increases the total cost through vaccination costs and adverse events among the *N*_*ct*_*/*2 vaccinated individuals. After a trial, at time *T*_*v*_ + *T*_*ct*_, the best policy is to not vaccinate. Notice that we do not use clinical trial information here, and that policies “trial, optimal at *T*_*v*_ + *T*_*ct*_”, “CT0” and “CT95” are trivially equivalent.

**Table App-1.** Summary simulation results for different vaccination policies, with uncertain parameters set to their prior expected value. Costs in 10^9^ USD.

| Policy | Cost |  |  |  |
| --- | --- | --- | --- | --- |
|  | Infections | Vaccination | Adverse events | Total |
| No trial, no vaccination | 28.34 | 0 | 0 | 28.34 |
| No trial, vaccination at $T_v$ | 26.34 | 9.34 | 13.84 | 49.52 |
| Trial, no vaccination | 28.34 | $3.22 \times 10^{-3}$ | $4.92 \times 10^{-3}$ | 28.35 |
| Trial, vaccination at $T_v + T_{ct}$ | 27.84 | 9.11 | 13.49 | 50.43 |
| Trial, optimal at $T_v + T_{ct}$ | 28.34 | $3.22 \times 10^{-3}$ | $4.92 \times 10^{-3}$ | 28.35 |
| CT0 | 28.34 | $3.22 \times 10^{-3}$ | $4.92 \times 10^{-3}$ | 28.35 |
| CT95 | 28.34 | $3.22 \times 10^{-3}$ | $4.92 \times 10^{-3}$ | 28.35 |

## C RCT performance in the ROC plane

The performance of clinical trials can also be illustrated with receiver operating characteristic (ROC) curves. In Figure App-3, we display the performance obtained in the ROC plane by varying safety threshold 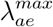, efficacy confidence level 1 − *α*, or both. The simulations are the same as in Figures 2, 3, and 4 respectively. The true positive rate is the proportion of simulations for which the vaccine passes both tests, among the simulations for which it is optimal to vaccinate at time *T*_*v*_ + *T*_*ct*_ following the trial. The false positive rate is the proportion of simulations for which the vaccine passes both tests, among the simulations for which it is optimal to not vaccinate at time *T*_*v*_ + *T*_*ct*_. Recall that the true positive and false positive rates of CT0 and CT95 can be retrieved from Table 3 as shown in Section 3.1. RCTs in the (0, 0) corner are too restrictive, RCTs in the (1, 1) corner are too permissive, and the (0, 1) corner corresponds to a perfect classifier. For some permissive safety thresholds, the RCT performs worse than random guess (dots below the line *y* = *x* in Figure (App-3d)).

**Figure App-3.**
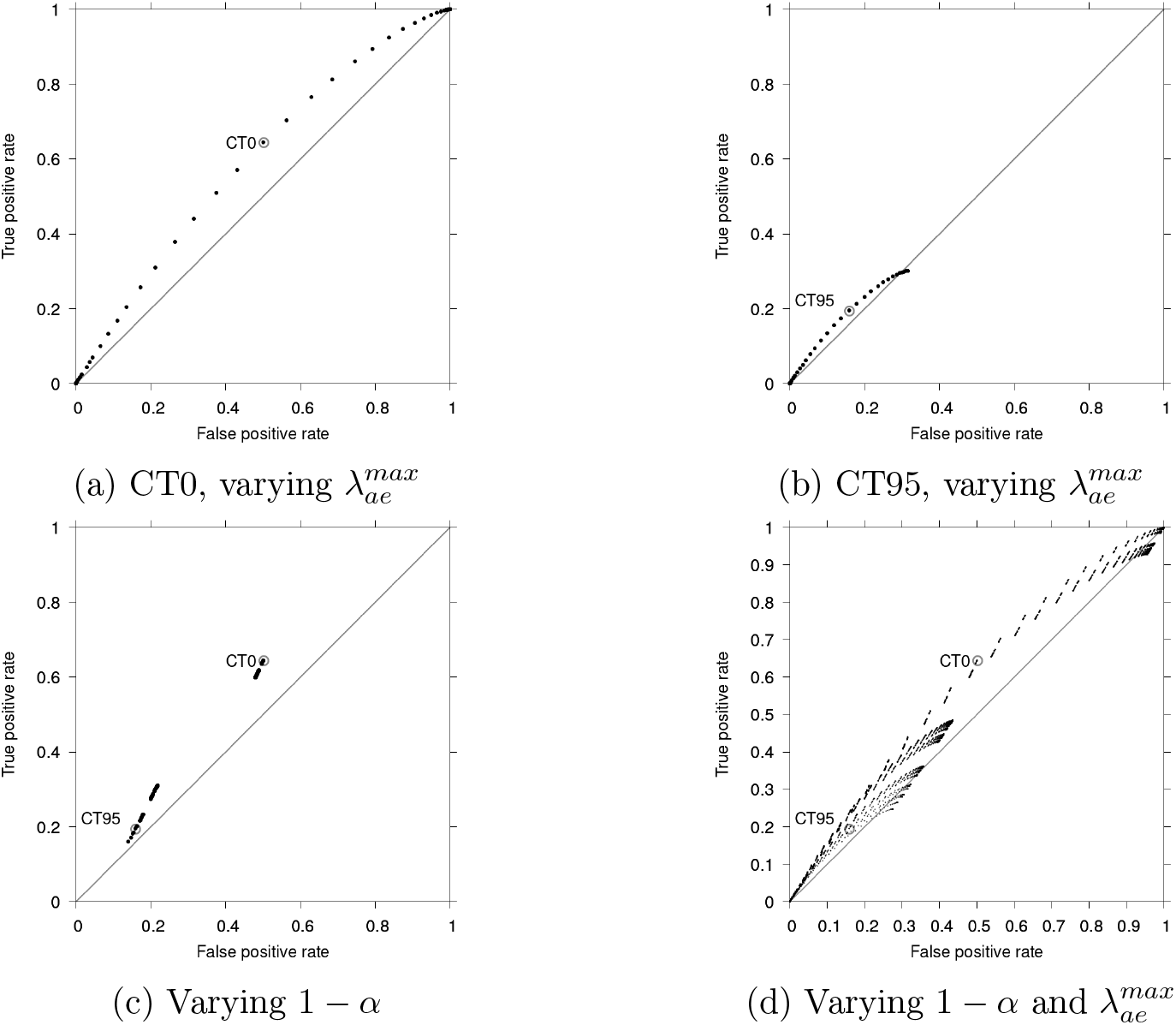
ROC curves obtained by varying safety threshold 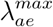, efficacy confidence level 1 − *α*, or both. Grey circles: CT0 and CT95.

## D Additional figure: varying *T*_*ct*_

**Figure App-4.**
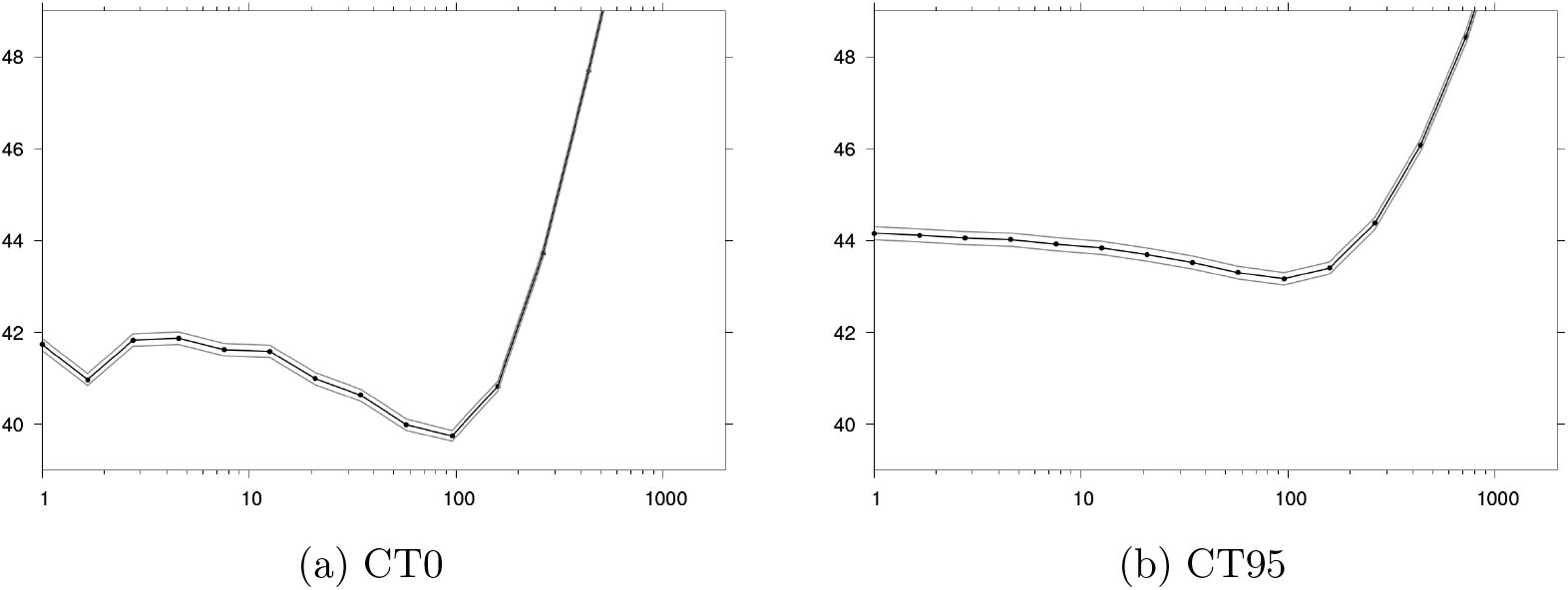
Total cost (10^9^ USD) as a function of the duration *T*_*ct*_ of the clinical trial (days) for policies “CT0” and “CT95”. Grey: 95% CI.

## Footnotes

1 Notice that this distinction is somewhat artificial. Think of the potential influence of the clinical trial (vaccinated individuals in the treatment arm) on the course of epidemic: it is an uncertain parameter of transmission that depends on trial random sampling.

2 But we note that it still seems to have influenced simulation-based vaccine trial design studies that typically assume an efficacy value, or consider a limited number of efficacy scenarios [35–42].

3 This issue remains the same irrespective of the considered summary estimand. For example, *effectiveness* estimands as presented e.g. in [11, 64] would likely have similar limitations. For efficacy testing, the present article looks primarily into incidence difference between RCT arms without loss of generality.

4 912.5 cases per year per 100,000 individuals.

5 Recall that we are using a deterministic transmission model.

1 It seems hard to consider that the entire population can be enrolled in a RCT.

